# Waist circumference thresholds predicting incident dysglycemia and type 2 diabetes in Black African men and women

**DOI:** 10.1101/2021.10.18.21265125

**Authors:** Julia H. Goedecke, Kim Nguyen, Clement Kufe, Maphoko Masemola, Tinashe Chikowore, Amy E. Mendham, Shane A. Norris, Nigel J. Crowther, Fredrik Karpe, Tommy Olsson, Andre Pascal Kengne, Lisa K Micklesfield

## Abstract

**Objective:** To determine the waist circumference (WC) thresholds for the prediction of incident dysglycemia and type 2 diabetes in Black South African (SA) men and women and compare these to advocated International Diabetes Federation (IDF) Europid thresholds.

**Research design and method:** In this prospective study, Black SA men (n=502) and women (n=527) from the Middle Aged Sowetan Cohort (MASC) study who had normal or impaired fasting glucose at baseline (2011-2015) were followed up through 2017-2018. Baseline measurements included anthropometry, blood pressure and fasting glucose, HDL-cholesterol and triglyceride concentrations. At follow-up, glucose tolerance was assessed using an oral glucose tolerance test. The Youden index was used to determine the optimal threshold of WC to predict incident dysglycemia and type 2 diabetes.

**Results:** In men, the optimal WC threshold was 96.8 cm for both dysglycemia and type 2 diabetes (sensitivity 56 and 70%, specificity 74 and 70%, respectively), which performed similarly to the IDF threshold of 94 cm. In women, the optimal WC threshold for incident dysglycemia was 91.8 cm (sensitivity 86%, specificity 37%) and for type 2 diabetes was 95.8 cm (sensitivity 85%, specificity 45%). In comparison, the IDF threshold of 80 cm in women had higher sensitivity (97 and 100%), but lower specificity (12 and 11%) to predict incident dysglycemia and type 2 diabetes, respectively.

**Conclusions:** In this first prospective study of incident type 2 diabetes in Africa, we show that African-specific WC thresholds perform better than the IDF WC thresholds to predict incident dysglycemia and type 2 diabetes.

## INTRODUCTION

The global prevalence of type 2 diabetes is increasing, with sub-Saharan Africa (SSA) having the highest projected relative rates of increase (1). The burden of type 2 diabetes in SSA is reflected by the high estimated type 2 diabetes-associated deaths (∼312,000 deaths in 2017), with 73% of these being in people under the age of 60 years, a higher proportion than any other region in the world (1). Within SSA, South Africa (SA) has the highest number of people with type 2 diabetes (1), and type 2 diabetes was the second leading cause of death in SA in 2016 (5.5% of deaths), and the highest amongst women (7.2% of deaths) (2). Notably, SSA has the highest proportion (59.7%) of people with undiagnosed type 2 diabetes (1). Accordingly, risk stratification that is accessible and cost effective is essential for the early detection of type 2 diabetes to prevent or delay the progression of the disease.

Obesity, in particular central obesity, are important risk factors for type 2 diabetes (3; 4). Although imaging techniques are more accurate measures of total and central adiposity, they are not practical or affordable for routine practice and population-based risk stratification. Accordingly, anthropometric measures are used as surrogate markers for risk stratification for type 2 diabetes. Body mass index (BMI) is the most commonly used proxy of total adiposity, while waist circumference (WC), waist-to-hip ratio (WHR) and waist-to-height ratio (WHtR) are used as proxy measures of central adiposity (5). Waist circumference requires only a single measure, is not reliant on a ratio that is difficult to interpret and is the most accepted marker of central adiposity and disease risk (4; 6).

Waist circumference represents the sum of abdominal visceral (VAT) and subcutaneous adipose tissue (SAT), with VAT being the most significant determinant of type 2 diabetes (7; 8). However, we and others have shown that for the same level of WC, Black Africans have less VAT than their white European counterparts (9-11). Accordingly, the WC threshold used for defining risk for type 2 diabetes may differ in Black Africans. Indeed, both the World Health Organization (WHO) and the International Diabetes Federation (IDF) acknowledge that optimal thresholds for abdominal obesity vary across different ethnicities and population groups (6; 12). Although, several studies in SSA have been undertaken to identify WC thresholds for risk, these have all been cross-sectional and relied on metabolic syndrome (MetS) (excluding WC) as the outcome (13-16). As there is no consensus on an appropriate WC threshold for Black Africans, the IDF has recommended the use of Europid thresholds (≥80 cm in women and ≥94 cm in men) for SSA (6). Prospective studies are therefore required to identify the optimal WC thresholds that identifies incident type 2 diabetes in Black African men and women.

While WC is regarded as a useful primary screening tool for type 2 diabetes, it is also a key feature of the MetS, which is also typically used in risk prediction for type 2 diabetes and cardiovascular diseases (6). The MetS is a cluster of risk factors that occur together more often than by chance alone, and in addition to WC, include elevated blood pressure, fasting glucose and triglycerides, and low fasting HDL-cholesterol concentrations (6). However, it is not clear whether including these additional MetS risk factors improves the discriminatory ability to predict type 2 diabetes in African men and women when compared to WC alone.

Therefore, the aim of the study was to determine the WC thresholds for the prediction of incident dysglycemia (prediabetes and type 2 diabetes) and type 2 diabetes in Black SA men and women, and to compare these to advocated Europid thresholds, as defined by the IDF. A secondary aim was to determine if the derived WC thresholds for the prediction of incident dysglycemia and type 2 diabetes performed similarly to the MetS in Black SA men and women.

## METHODS

### Design, study population and setting

Baseline data collection was part of the African WITS-INDEPTH Partnerships for Genomic Research (AWI-Gen) Collaborative Centre which is a Human Heredity and Health in Africa (<u>H3A</u>) Consortium study (17; 18), and included 1027 men and 1004 women from which the Middle-aged Soweto Cohort (MASC) was randomly selected (n=1112). The MASC cohort is a longitudinal study of Black SA men and women residing in Soweto, SA on whom baseline data was collected between 2011 and 2015, and again between January 2017 and August 2018 (Figure 1). Data in this study was collected from a sample of 1029 participants (502 men and 527 women) who were representative of the AWI-Gen sample and did not differ in terms of age, sex, sociodemographic or lifestyle factors from the main cohort (data not shown). Only participants with normal glucose tolerance or impaired fasting glucose, and anthropometric measures at baseline, as well as measures of glycaemia from an oral glucose tolerance test at follow-up, were included in this analysis (Figure 1). Complete data was available for 890 participants (452 men and 438 women).

**Figure 1.**
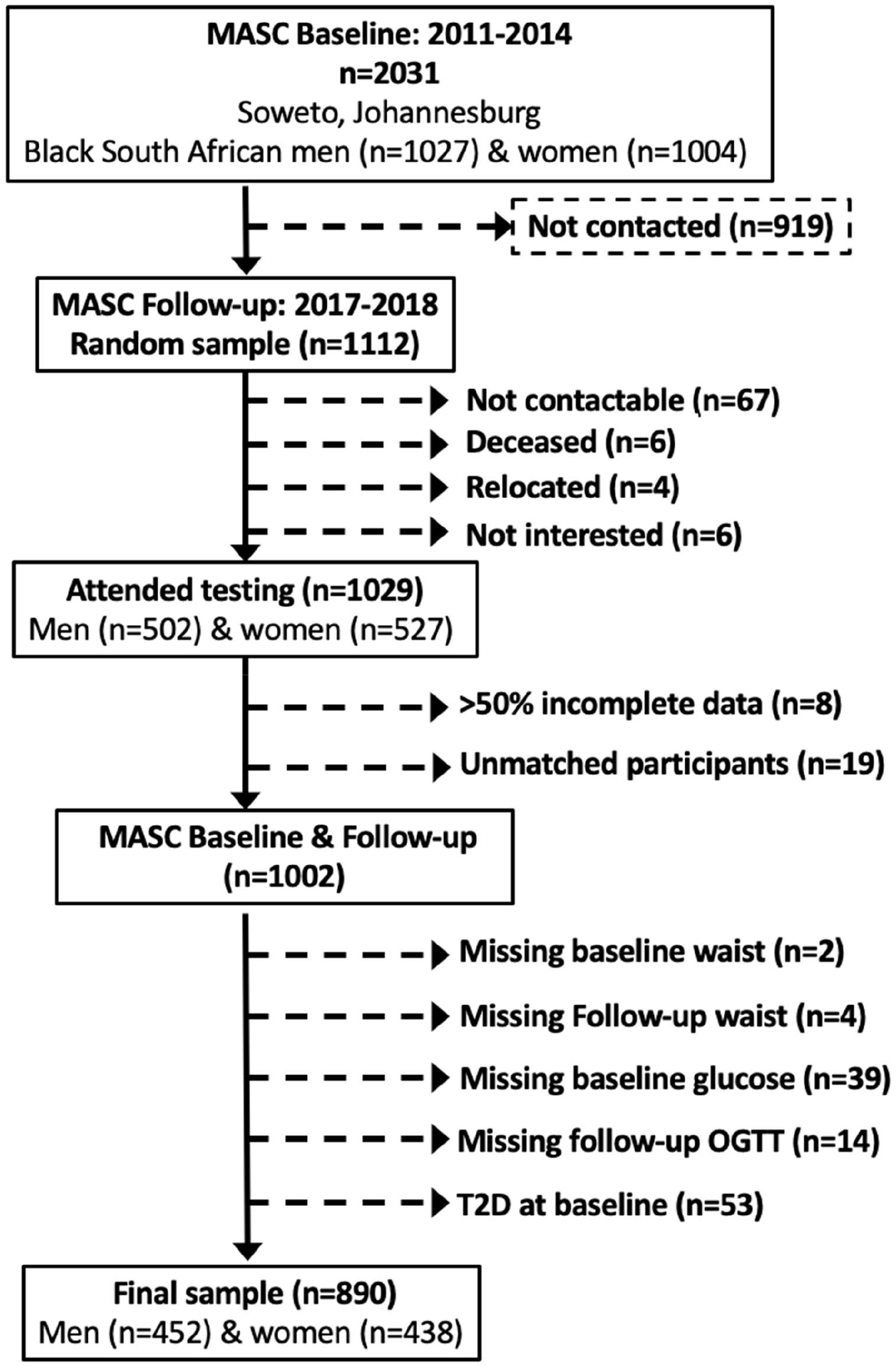
Consort Diagram for the Middle Aged Sowetan Cohort (MASC) waist circumference study

The study was conducted in accordance with the Declaration of Helsinki and was approved by the Human Research Ethics Committee (HREC) (Medical) of the University of the Witwatersrand (Clearance Certificate No. M160604 and M160975). Prior to inclusion in the study, all the procedures and possible risks were explained to the participants prior to providing signed consent.

### Socio–demographic and health questionnaires

The same interviewer-administered questionnaire was completed at both time points and included age, current employment (employed/not employed), and highest educational level attained (no formal schooling/elementary school, secondary school, tertiary education). In addition, participants were asked to bring all other current medications, including diabetes medications, to the testing facility for recording and verification. Participants were classified as current smokers/non-smokers and current alcohol consumers/non-consumers.

### Anthropometry and blood pressure

Weight was measured to the nearest 0.1 kg using a TANITA digital scale (model: TBF-410, TANITA Corporation, US). Height was measured to the nearest 0.1 cm using a wall–mounted stadiometer (Holtain, Crymych, UK). Waist circumference and hip circumference were measured to the nearest 0.1 cm with a non–stretchable tape. Waist circumference was measured in the mid–axillary line at the midpoint between the lower margin of the last palpable rib and the top of the iliac crest at the end of normal expiration, and hip circumference was measured as the greatest protrusion of the buttocks (12). Waist-to-hip ratio (WHR), waist-height ratio (WHtR) and body mass index (BMI) were calculated. Participants were categorized according to the WHO criteria: underweight (BMI <18.5 kg/m^2^), normal weight (BMI: 18.5-24.9 kg/m^2^), overweight (25-29.9 kg/ m^2^) and obese (BMI ≥ 30 kg/ m^2^).

Systolic and diastolic blood pressure were measured on the left arm using a digital blood pressure monitor (Omron M6, Kyoto, Japan) and appropriate cuffs. After the participant had been seated for at least five minutes, three blood pressure readings were taken at two-minute intervals. For each participant, the average of the second and third readings were used in the analyses.

### Blood sampling and biochemistry

At both baseline and follow-up, blood samples were drawn after an overnight fast (10-12 hours) for the measurement of plasma glucose and serum lipid (total cholesterol, HDL-cholesterol, LDL-cholesterol and triglyceride) concentrations. In the follow-up sample only, participants then completed a standard 2–hour oral glucose tolerance test (OGTT). After ingestion of 75 g of anhydrous glucose in 250 ml water within 5 minutes, blood samples were drawn at 30 min intervals for 2 hours for the subsequent determination of plasma glucose concentrations. Participants with known diabetes and/or those with fasting blood glucose ≥11.1 mmol/l (ACCU-CHEK^®^ point-of-care glucose analyzer, MedNet GmbH, Munster, Germany) did not complete the OGTT.

Serum lipid concentrations at baseline and plasma glucose concentrations at baseline and follow-up were measured on the Randox RX Daytona Chemistry Analyzer using enzymatic methods (Randox Laboratories Ltd., Crumlin, Northern Ireland). The LDL-cholesterol concentrations were calculated using the Friedewald equation (19).

### Glucose tolerance and metabolic syndrome (MetS) classification

Glucose tolerance was defined based on the WHO criteria (20). At baseline, only fasting glucose samples were available, and hence the participants were classified as having normal glucose tolerance (NGT) if fasting glucose was <6.1 mmol/L; impaired fasting glucose (IFG) if fasting glucose was 6.1-6.9 mmol/l; or type 2 diabetes if fasting glucose was ≥7 mmol/l and/or taking type 2 diabetes medications. Only those with NGT or IFG at baseline were included in this study. At follow-up, glucose tolerance was defined based on both fasting and 2 hour OGTT results as follows: NGT if fasting glucose was <6.1 mmol/l and 2-hour post glucose load was <7.8 mmol/L; IFG (as defined above); impaired glucose tolerance (IGT) if 2-hour post glucose load was 7.8– 11.0 mmol/L; and type 2 diabetes if fasting glucose was >7.0 mmol/L and/or 2-hour post glucose load ≥11.1 mmol/L. Participants who were taking diabetes medications were classified with type 2 diabetes. At follow-up dysglycemia, which encompasses both prediabetes and diabetes, was defined as IFG and/or IGT and/or type 2 diabetes.

The presence of the MetS was based on the 2009 harmonized criteria (6). Participants with three or more of the following components were classified as having the MetS: i) elevated waist circumference (≥94 cm in men or ≥80 cm in women); ii) elevated fasting triglycerides (≥1.7 mmol/L); iii) reduced fasting HDL cholesterol (<1.0 mmol/L in men or <1.3 mmol/L in women); iv) elevated blood pressure (≥130 mmHg for systolic and/or ≥85 mmHg for diastolic and/or using blood pressure medication); v) elevated fasting glucose (≥5.6 mmol/L and/or using diabetes medication).

### Statistics

Data analysis was conducted in STATA SE Version 15 (StataCorp, College Station, Tx, USA). Normality of the data was assessed using Shapiro Wilks test. As all the descriptive variables were skewed, the continuous variables are presented as median (25-75^th^ percentiles) and the categorical variables are presented as frequencies and percentages. Wilcoxon-Mann-Whitney tests and Chi-square tests were used to compare continuous and categorical variables between men and women, respectively. Receiver operating characteristic (ROC) area-under-the-curves (AUC) were used to assess and compare the ability of the baseline WC and other anthropometric measures to predict incident dysglycemia (prediabetes or type 2 diabetes) and type 2 diabetes at follow-up. For the prediction of incident dysglycemia, only those with NGT at baseline were included in the analysis, whereas for the prediction of incident type 2 diabetes, those with NGT and IFG at baseline were included in the analysis. Optimal WC thresholds to predict incident dysglycemia and type 2 diabetes were determined using Youden’s index in men and women separately (21). The prognostic performance of the WC thresholds derived in this longitudinal study were assessed alongside the IDF-defined threshold, as well as WC thresholds defined in other South African and African cross-sectional studies that have been used to predict the presence of at least two components of the MetS, excluding WC (22). These studies were used as comparators as, to our knowledge, there are no other studies that have previously defined WC thresholds for predicting incident type 2 diabetes in Africa. Finally, we determined whether including additional MetS risk factors together with the derived WC threshold improved the prediction of incident dysglycemia and type 2 diabetes compared to the derived WC thresholds alone.

## RESULTS

### Participant characteristics

At baseline, the sample were middle-aged (∼50 years) with men being slightly older than women (Table 1). More men were employed (65.0% vs. 53.0%, p<0.001) and had completed secondary (75.0 vs. 16.9%) and tertiary education (16.4% vs. 0%), compared to women (both p<0.001). In addition, more men than women currently smoked any form of tobacco (52.2% vs. 5.3%, p<0.001). In contrast, women had significantly higher BMI and a greater proportion of women compared to men were classified with overweight or obesity (88.6% vs. 47.4%, p<0.001, Table 1). Accordingly, WC and WHtR were higher, but WHR was lower in women compared to men (Table 1). Fasting glucose concentrations were higher in men compared to women, but women had higher triglyceride, total- and LDL-cholesterol concentrations compared to men, while HDL-cholesterol concentrations did not differ (Table 1). Similarly, both systolic and diastolic blood pressure did not differ by sex. In terms of the sex differences in MetS criteria, a greater proportion of women had elevated WC, while a greater proportion of men had elevated fasting glucose and reduced HDL-cholesterol concentrations; however, the prevalence of MetS did not differ significantly between men and women.

**Table 1.** Characteristics of men and women at baseline

|  | Men (n=452) | Women (n=438) | P value |
| --- | --- | --- | --- |
| Age (years) | 50 (45-55) | 49 (45-54) | 0.017 |
| Anthropometry |  |  |  |
| BMI (kg/m2) | 24.5 (20.8-29.1) | 32.8 (28.5-37.3) | <0.001 |
| Waist circumference (cm) | 88.1 (78.0-100.0) | 98.1 (89.5-107.0) | <0.001 |
| Hip circumference (cm) | 99.4 (91.2-106.1) | 117.0 (108.5-126.0) | <0.001 |
| WHR | 0.90 (0.85-0.95) | 0.84 (0.78-0.89) | <0.001 |
| WHtR | 0.52 (0.46-0.58) | 0.62 (0.56-0.67) | <0.001 |
| BMI categories |  |  |  |
| Underweight (%(n)) | 10.0 (46) | 0.5 (2) | <0.001 |
| Normal weight (%(n)) | 42.6 (196) | 11.0 (48) |  |
| Overweight (%(n)) | 26.5 (122) | 21.9 (96) |  |
| Obese (%(n)) | 20.9 (96) | 66.7 (292) |  |
| Biochemistry |  |  |  |
| Fasting glucose (mmol/l) | 5.1 (4.7-5.4) | 4.8 (4.5-5.2) | <0.001 |
| Triglycerides (mmol/l) | 0.9 (0.6-1.3) | 1.0 (0.7-1.4) | <0.001 |
| Total cholesterol (mmol/l) | 4.1 (3.4-4.8) | 4.5 (3.8-5.2) | <0.001 |
| HDL-cholesterol (mmol/l) | 1.2 (0.9-1.5) | 1.2 (1.0-1.5) | 0.063 |
| LDL-cholesterol (mmol/l) | 2.5 (1.8-3.1) | 2.7 (2.2-3.3) | <0.001 |
| Blood pressure |  |  |  |
| Systolic (mmHg) | 129.0 (117.0-144.0) | 128.5 (117.0-141.5) | 0.518 |
| Diastolic (mmHg) | 88.8 (79.5-96.5) | 87.5 (79.0-96.0) | 0.357 |
| Metabolic syndrome (JIS) |  |  |  |
| Elevated WC (%(n)) | 37.6 (170) | 90.4 (396) | <0.001 |
| Elevated fasting glucose (%(n)) | 15.9 (72) | 10.5 (46) | 0.017 |
| Elevated triglycerides (%(n)) | 13.1 (59) | 14.4 (63) | 0.564 |
| Reduced HDL-cholesterol (%(n)) | 69.3 (313) | 46.1 (202) | <0.001 |
| Elevated blood pressure (%(n)) | 61.3 (279) | 59.6 (261) | 0.514 |
| MetS (%(n)) | 30.3 (137) | 35.4 (155) | 0.107 |

Values are median (25-75<sup>th</sup> percentile). WHR, waist-to-hip ratio; WHtR, waist-to-height ratio; MetS, metabolic syndrome, defined using the Joint Interim Statement (JIS) (6) and defined as meeting any three of the following criteria: elevated waist circumference ( $\geq 80$ cm in women; $\geq 94$ cm in men); elevated blood glucose ( $\geq 5.6$ mmol/l and/or using diabetes medication); elevated fasting triglycerides ( $\geq 1.7$ mmol/L); reduced fasting HDL-cholesterol ( $<1.0$ mmol/L in men, $<1.3$ mmol/L in women); elevated blood pressure ( $\geq 135/85$ mmHg and/or using antihypertension medication).

The median (25-75^th^ percentile) follow-up times were 3.1 (2.9-3.5) years in men and 4.8 (4.0-5.5) years in women. Of the 430 men and 421 women with NGT at baseline, 73 men and 101 women developed dysglycemia at follow-up, resulting in a cumulative incidence (95% confidence incidence (CI)) of 17.0 (13.4-28.1)% in men and 24.0 (19.9-41.1)% in women. Of the 452 men and 438 women with NGT or IFG at baseline, 20 men and 47 women developed type 2 diabetes at follow-up, resulting in a lower cumulative incidence of type 2 diabetes (95% CI) in men (4.4 (2.5-5.9)%) compared to women (10.7 (7.8-16.8)%).

### Comparative abilities of anthropometric measures to predict incident dysglycemia and type 2 diabetes

When comparing the discriminatory power of the anthropometric measures to predict incident dysglycemia and type 2 diabetes in men and women (Supplementary Table S1), we showed that there were no significant differences between any of the AUC’s, except where WHtR performed better than BMI in predicting dysglycemia in men (p=0.017). As WC is a single simple measure that is most commonly used for risk prediction (6) and recommended for clinical practice (4), we chose to use this measure in all subsequent analyses.

### Performance of different WC thresholds to predict incident dysglycemia and type 2 diabetes

The ROC analyses for WC to predict incident dysglycemia and type 2 diabetes in men and women are presented in Figure 2. Waist circumference showed acceptable discrimination to predict dysglycemia and type 2 diabetes in men and women, with the AUCs being higher in men than women (Figure 2). Based on the Youden’s index, the optimal WC thresholds to predict incident dysglycemia in men and women were 96.8 cm and 91.8 cm, respectively (Table 2). In men, this threshold was similar to the IDF threshold of 94 cm, and accordingly had similar sensitivity, specificity, and positive and negative predictive values. However, the threshold of 96.8 cm was higher than those derived from cross-sectional studies of other African populations to detect MetS (84-90 cm), with a resultant lower sensitivity, but higher specificity.

**Figure 2.**
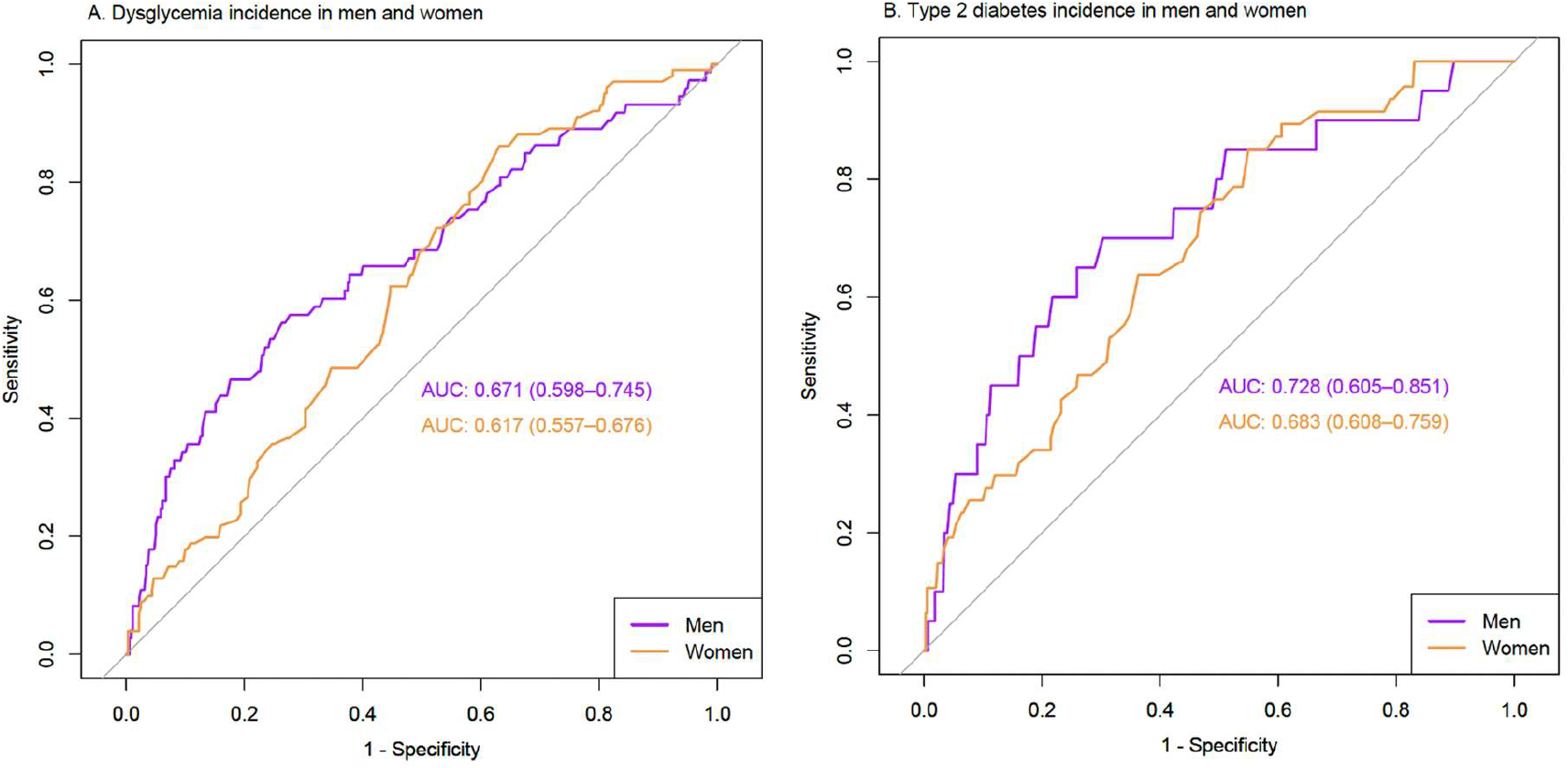
Receiver operating characteristics (ROC) curve of waist circumference to predict incident dysglycemia (A) and type 2 diabetes (B) in Black African men and women.

**Table 2.** Performance measures of different waist circumference thresholds to predict incident dysglycemia in Black African men and women

| Reference | Cut-off (cm) | Sensitivity (95%CI) | Specificity (95%CI) | PPV (95%CI) | NPV (95%CI) | Youden index (95% CI) |
| --- | --- | --- | --- | --- | --- | --- |
| **Men** |  |  |  |  |  |  |
| Youden | 96.8 | 0.56<br>(0.44-0.68) | 0.74<br>(0.69-0.78) | 0.30<br>(0.23-0.39) | 0.89<br>(0.85-0.92) | 0.30<br>(0.13-0.46) |
| IDF (6) | 94 | 0.59<br>(0.47-0.70) | 0.68<br>(0.63-0.73) | 0.27<br>(0.21-0.35) | 0.89<br>(0.85-0.92) | 0.27<br>(0.10-0.43) |
| Matsha et al. (13) | 90 | 0.66<br>(0.54-0.76) | 0.59<br>(0.54-0.64) | 0.25<br>(0.19-0.31) | 0.89<br>(0.85-0.93) | 0.25<br>(0.07-0.40) |
| Ekoru et al. (15) | 81 | 0.82<br>(0.72-0.90) | 0.35<br>(0.30-0.40) | 0.20<br>(0.16-0.26) | 0.91<br>(0.84-0.95) | 0.17<br>(0.01-0.30) |
| Motala et al. (16) | 86 | 0.68<br>(0.57-0.79) | 0.48<br>(0.43-0.53) | 0.21<br>(0.16-0.27) | 0.88<br>(0.83-0.92) | 0.17<br>(0.01-0.32) |
| Peer et al. (23) | 83.9 | 0.74<br>(0.62-0.84) | 0.45<br>(0.40- 0.50) | 0.22<br>(0.17-0.27) | 0.89<br>(0.84-0.94) | 0.19<br>(0.02-0.34) |
| **MetS** | 96 | 0.44<br>(0.32-0.56) | 0.77<br>(0.72-0.81) | 0.28<br>(0.20-0.37) | 0.87<br>(0.83-0.90) | 0.21<br>(0.04-0.37) |
| **Women** |  |  |  |  |  |  |
| Youden | 91.8 | 0.86<br>(0.78-0.92) | 0.37<br>(0.32-0.42) | 0.30<br>(0.25-0.36) | 0.89<br>(0.83-0.94) | 0.23<br>(0.09-0.35) |
| IDF (6) | 80 | 0.97<br>(0.92-0.99) | 0.12<br>(0.09-0.16) | 0.26<br>(0.22-0.31) | 0.93<br>(0.81-0.99) | 0.09<br>(0.01-0.16) |
| Matsha et al. (13) | 90 | 0.88<br>(0.80-0.94) | 0.31<br>(0.26- 0.36) | 0.29<br>(0.24-0.34) | 0.89<br>(0.82-0.94) | 0.19<br>(0.06-0.30) |
| Ekoru et al. (15) | 81 | 0.97<br>(0.92-0.99) | 0.14<br>(0.10- 0.18) | 0.26<br>(0.22-0.31) | 0.94<br>(0.83-0.99) | 0.11<br>(0.02-0.18) |
| Motala et al. (16) | 92 | 0.86<br>(0.78-0.92) | 0.37<br>(0.32- 0.42) | 0.30<br>(0.25-0.36) | 0.89<br>(0.83-0.94) | 0.23<br>(0.09-0.35) |
| Crowther et al. (14) | 91.5 | 0.86<br>(0.78-0.92) | 0.36<br>(0.30- 0.41) | 0.30<br>(0.25-0.35) | 0.89<br>(0.82-0.94) | 0.22<br>(0.08-0.33) |
| Peer et al. (23) | 94 | 0.76<br>(0.67-0.84) | 0.43<br>(0.37-0.48) | 0.30<br>(0.24-0.36) | 0.85<br>(0.79-0.90) | 0.19<br>(0.04-0.33) |
| **MetS** | 96 | 0.38<br>(0.28-0.48) | 0.80<br>(0.75-0.84) | 0.37<br>(0.28-0.47) | 0.80<br>(0.75-0.84) | 0.17<br>(0.03-0.32) |

Dysglycemia defined as impaired fasting glucose and/or impaired glucose tolerance or type 2 diabetes; IDF, International Diabetes Federation; Sensitivity = TP/(TP+FN); Specificity = TN/(TN+FP); PPV, positive predictive value = TP/(TP+FP); NPV, negative predictive value = sensitivity/1-specificity; Youden's index = (sensitivity + specificity)– 1, where TP, true positive; FP, false positive; TN, true negative; FN, false negative; MetS, metabolic syndrome using a waist circumference threshold of 96 cm for men and women derived from this study.

In women, the threshold of 91.8 cm to predict incident dysglycemia was higher than the IDF recommended threshold of 80 cm but was similar to most thresholds from other African studies to detect MetS (Table 2). Although the sensitivity was lower than the IDF threshold (0.86 vs. 0.97), the specificity (0.37 vs. 0.12) was higher using the derived threshold of 91.8 cm.

The optimal WC threshold to predict incident type 2 diabetes in men (Table 3) was the same as that for dysglycemia (96.8 cm) and consequently all the performance variables are the same as those reported for dysglycemia in Table 2. In contrast, the optimal WC threshold to predict incident type 2 diabetes in women was 95.8 cm (Table 3), which is similar to the threshold for dysglycemia and type 2 diabetes in men, but higher than the threshold for dysglycemia (91.8 cm), and the IDF cut point (80 cm) and most other thresholds derived to detect MetS in African women (81-94 cm). Although the sensitivity of the optimal threshold of 95.8 cm was lower than IDF (0.85 vs. 1.00) and other African studies (0.87-1.00), the specificity was higher (0.45 vs. 0.11 and 0.12-0.40.

**Table 3.** Performance measures of different waist circumference thresholds to predict incident type 2 diabetes in Black African men and women

| Reference | Cut-off (cm) | Sensitivity (95%CI) | Specificity (95%CI) | PPV (95%CI) | NPV (95%CI) | Youden index (95% CI) |
| --- | --- | --- | --- | --- | --- | --- |
| **Men** |  |  |  |  |  |  |
| Youden | 96.8 | 0.70<br>(0.46-0.88) | 0.70<br>(0.65-0.74) | 0.10<br>(0.05-0.16) | 0.98<br>(0.96- 0.99) | 0.40<br>(0.11- 0.62) |
| IDF (6) | 94 | 0.70<br>(0.46-0.88) | 0.64<br>(0.59-0.68) | 0.08<br>(0.05-0.13) | 0.98<br>(0.95- 0.99) | 0.34<br>(0.05-0.57) |
| Matsha et al. (13) | 90 | 0.75<br>(0.51-0.91) | 0.55<br>(0.50-0.59) | 0.07<br>(0.04-0.11) | 0.98<br>(0.95- 0.99) | 0.30<br>(0.01-0.51) |
| Ekoru et al. (15) | 81 | 0.90<br>(0.68-0.99) | 0.32<br>(0.27-0.36) | 0.06<br>(0.03-0.09) | 0.99<br>(0.95- 1.00) | 0.22<br>(-0.04-0.35) |
| Motala et al. (16) | 86 | 0.85<br>(0.62-0.97) | 0.45<br>(0.40-0.50) | 0.07<br>(0.04-0.10) | 0.98<br>(0.96- 1.00) | 0.30<br>(0.02-0.46) |
| Peer et al. (23) | 83.9 | 0.85<br>(0.62-0.87) | 0.41<br>(0.37-0.46) | 0.06<br>(0.04-0.10) | 0.98<br>(0.95- 1.00) | 0.26<br>(-0.01-0.43) |
| **MetS** | 96 | 0.45<br>(0.23-0.68) | 0.72<br>(0.68-0.77) | 0.07<br>(0.03-0.13) | 0.97<br>(0.94-0.98) | 0.17<br>(-0.09-0.45) |
| **Women** |  |  |  |  |  |  |
| Youden | 95.8 | 0.85<br>(0.72-0.94) | 0.45<br>(0.40-0.50) | 0.16<br>(0.11-0.21) | 0.96<br>(0.92 0.98) | 0.30<br>(0.12- 0.44) |
| IDF (6) | 80 | 1.00<br>(0.92-1.00) | 0.11<br>(0.08-0.14) | 0.12<br>(0.09-0.15) | 1.00<br>(0.92- 1.00) | 0.11<br>(0.01-0.14) |
| Matsha et al. (13) | 90 | 0.91<br>(0.80-0.98) | 0.28<br>(0.23-0.32) | 0.13<br>(0.10-0.17) | 0.96<br>(0.91- 0.99) | 0.19<br>(0.03-0.30) |
| Ekoru et al. (15) | 81 | 1.00<br>(0.92-1.00) | 0.12<br>(0.09-0.16) | 0.12<br>(0.09-0.16) | 1.00<br>(0.93- 1.00) | 0.12<br>(0.02-0.16) |
| Motala et al. (16) | 92 | 0.91<br>(0.80-0.98) | 0.33<br>(0.29-0.38) | 0.14<br>(0.10-0.19) | 0.97<br>(0.93- 0.99) | 0.24<br>(0.08-0.36) |
| Crowther et al. (14) | 91.5 | 0.91<br>(0.80-0.98) | 0.32<br>(0.28-0.37) | 0.14<br>(0.10-0.18) | 0.97<br>(0.92- 0.99) | 0.24<br>(0.07-0.35) |
| Peer et al. (23) | 94 | 0.87<br>(0.74-0.95) | 0.40<br>(0.36-0.45) | 0.15<br>(0.11-0.20) | 0.96<br>(0.92-0.99) | 0.28<br>(0.10-0.41) |
| **MetS** | 96 | 0.49<br>(0.34-0.64) | 0.76<br>(0.72-0.80) | 0.20<br>(0.13-0.28) | 0.93<br>(0.89-0.95) | 0.25<br>(0.06-0.44) |

IDF, International Diabetes Federation; Sensitivity = $TP/(TP+FN)$ ; Specificity = $TN/(TN+FP)$ ; PPV, positive predictive value = $TP/(TP+FP)$ ; NPV, negative predictive value = $sensitivity/1-specificity$ ; Youden's index = $(sensitivity + specificity) - 1$ , where TP, true positive; FP, false positive; TN, true negative; FN, false negative; MetS, metabolic syndrome using a waist circumference threshold of 96 cm for men and women derived from this study.

### Comparative ability of derived WC thresholds vs. MetS to predict incident dysglycemia and type 2 diabetes

We then determined whether including additional MetS risk factors together with the derived WC thresholds improved the prediction of incident dysglycemia and type 2 diabetes compared to the derived WC thresholds alone. In Tables 2 and 3, we showed that the cut-point to predict incident dysglycemia and T2D in men, and T2D in women were similar (∼96 cm). Other African studies that have examined cut-points to detect MetS have also suggested similar cut-points for men and women (13; 15; 16). Thus, we used the WC cut-point of 96 cm in both men and women as the WC component of MetS and compared this to the WC cut-points alone, to predict incident dysglycemia and T2D in men and women (Tables 2 and 3). Despite including additional risk factors, MetS had lower sensitivity, but similar specificity compared to the optimal WC threshold of 96.8 cm to predict incident dysglycemia and type 2 diabetes in men (Tables 2 and 3). In contrast, the sensitivity of MetS using a WC threshold of 96cm to predict both incident dysglycemia and type 2 diabetes in women was lower than the derived thresholds alone, while the specificity was higher (Tables 2 and 3).

## DISCUSSION

This is the first prospective study to examine WC thresholds to predict incident dysglycemia and type 2 diabetes in an African population and showed that the optimal thresholds differed to those in European populations. The optimal thresholds to predict incident dysglycemia and type 2 diabetes in Black SA men were 96.8 cm for both outcomes and in women 91.8 and 95.8 cm, respectively. Importantly, these African-specific WC thresholds had higher specificity than the IDF Europid thresholds.

The WC thresholds to predict incident dysglycemia and type 2 diabetes in women were higher than the IDF threshold of 80 cm. This is consistent with the findings of several cross-sectional studies in SA that detected MetS, defined as at least two components of the MetS excluding WC, and reported optimal thresholds of 90-94 cm (13; 14; 16; 23). We showed that in women the thresholds of 91.8 and 95.8 cm had lower sensitivity and higher specificity than the IDF threshold to predict incident dysglycemia and type 2 diabetes, respectively. The specificity of the IDF threshold was as low as 0.11, suggesting 89% of Black SA women who will remain free of dysglycemia or type 2 diabetes over time may be incorrectly classified among those who will go on to develop the conditions if this threshold was used alone as a risk screening tool.

Nonetheless, the devised African-specific threshold still had low specificity (0.37 and 0.45) for incident dysglycemia and type 2 diabetes in Black SA women. This was also lower than that reported in Black SA men using the derived threshold of 96.8 cm (specificity ∼0.70). The poor discriminatory ability of the WC thresholds to predict incident dysglycaemia and type 2 diabetes in women may be due to their high levels of obesity (66.7% vs. 20.9%) and central obesity (90% WC >80 cm vs. 37.6% WC >94 cm) compared to the men in this sample. However, this is representative of men and women of this age group in SA (24). Further, these discrepant findings may be explained by the stronger association between total and central adiposity and type 2 diabetes risk in Black SA men compared to Black SA women (Kufe et al, in review). Waist circumference incorporates both VAT and SAT; and it has been shown in Black South Africans and African-born Black people living in America that for the same WC, women have less VAT and more abdominal SAT than men (25) (Kufe et al., in review). Higher SAT for every level of VAT explained the higher WC cut point required for predicting insulin resistance in African-born black women living in America compared to their male counterparts (96 cm vs. 91 cm) (25). Further, a prospective study in Black SA women showed that only VAT, but not SAT, predicted the development of T2D (7). Accordingly, high levels of SAT in Black SA women may mask the association with VAT and lead to poor discriminatory power of WC to predict incident type 2 diabetes. Previous studies from SSA have suggested that the WC of risk should be similar for African men and women (13; 15; 16). The findings from this study support this recommendation. Indeed, a WC of 96.8 cm predicted both incident dysglycemia and type 2 diabetes in men, whilst a very similar WC of 95.8 cm predicted incident type 2 diabetes in women. However, larger longitudinal studies are required to confirm our findings for the SA population. We do agree that WC thresholds are dependent on the underlying obesity prevalence and should be region specific (15).

Another important finding of this study was that when including additional MetS risk factors together with the derived WC threshold to predict incident dysglycemia and type 2 diabetes in men, the predictive accuracy did not change. In contrast, in women, the inclusion of the additional MetS risk factors resulted in a decrease in sensitivity (0.85-0.86 to 0.38-0.49), but an increase in specificity (0.37-0.45 to 0.80-0.76). In both men and women, the most common feature of MetS were reduced HDL-cholesterol and elevated blood pressure. In African populations, reduced HDL-cholesterol levels are not necessarily a marker of cardiometabolic risk, and the WHO sex-based cut-offs are inappropriate (26-28); while blood pressure is a risk factor for cardiovascular disease rather than type 2 diabetes. Further, it has been previously shown that MetS and its components, in particular triglyceride and HDL-cholesterol levels, are not associated with insulin resistance in Black African women (29-31) and that MetS may not be a good indicator of cardiometabolic risk in Black African populations (32-34). The time and cost associated with these additional measures is unlikely to offset the reduction in false positives associated with WC measures alone in Black SA women. Concomitantly, this highlights the need for future studies to establish accessible and cost-effective risk biomarkers that have high sensitivity and specificity for the early detection of type 2 diabetes in SSA.

The major strengths of this study are the prospective design and the diagnosis of incident dysglycemia and type 2 diabetes using an OGTT. To date, all studies that have explored thresholds for WC have been cross-sectional and were designed for identifying the optimal WC cut point for detecting MetS (13; 14; 16; 23). We used an OGTT to diagnose type 2 diabetes at follow-up, which is considered the gold standard, particularly in African populations where fasting glucose and HbA1c may perform sub-optimally (35-38). Another strength of this study is the inclusion of equal numbers of men and women. Most studies in SA have either focused on women only or included small samples of men (13-16; 23). Limitations of the study include the relatively small sample size that precluded the validation of the threshold in a sub-sample of the participants.

In conclusion, we show for the first time using prospective cohort data from SA that African-specific thresholds derived for WC perform better than the IDF Europid WC thresholds for predicting incident dysglycemia and type 2 diabetes in Black SA men and women.

## Data Availability

Some or all datasets generated during and/or analyzed during the current study are not publicly available but are available from the corresponding author on reasonable request.

## Author contributions

The authors confirm contribution to the paper as follows: study conception and design: JHG and LKM; data collection: CK, MM, TC; data analysis: JHG, KN, APK; interpretation of results: JHG, KN, APK, AEM, SAM, NJC, FK, TO, LKM; draft manuscript preparation: JHG; All authors reviewed the results and approved the final version of the manuscript.

Julia H. Goedecke is the guarantor and takes full responsibility for the contents of the manuscript.

## Acknowledgements

We are grateful to the participants as well as the following DPHRU staff for their input during data collection and entry: Vukosi Mkansi, Sphume Thango, Mosadiapula Nakedi, Bonisiwe Mlambo, Melikhanya Soboyisi, Tshifhiwa Ratshikombo, and team.

---

**Supplementary Table S1.** Comparative abilities of anthropometric measures at baseline to predict incident dysglycemia and type 2 diabetes at follow-up in Black African men and women

|  | AUC (95%CI) | P value vs. WC | P value vs. WHR | P value vs. WHtR |
| --- | --- | --- | --- | --- |
| **Dysglycemia** |  |  |  |  |
| **Men (n=429)** |  |  |  |  |
| Waist circumference | 0.674 (0.600-0.748) | - |  |  |
| Waist-to-hip ratio | 0.655 (0.584-0.726) | 0.591 | - |  |
| Waist-to-height ratio | 0.681 (0.610-0.751) | 0.425 | 0.395 | - |
| Body mass index | 0.653 (0.578-0.727) | 0.073 | 0.938 | **0.017** |
| **Women (n=420)** |  |  |  |  |
| Waist circumference | 0.620 (0.561-0.679) | - |  |  |
| Waist-to-hip ratio | 0.625 (0.561-0.688) | 0.866 | - |  |
| Waist-to-height ratio | 0.639 (0.579-0.699) | 0.058 | 0.593 | - |
| Body mass index | 0.610 (0.550-0.670) | 0.623 | 0.731 | 0.142 |
| **Type 2 diabetes** |  |  |  |  |
| **Men (n=451)** |  |  |  |  |
| Waist circumference | 0.728 (0.605-0.851) | - |  |  |
| Waist-to-hip ratio | 0.710 (0.604-0.816) | 0.587 | - |  |
| Waist-to-height ratio | 0.724 (0.603-0.845) | 0.686 | 0.683 | - |
| Body mass index | 0.688 (0.556-0.821) | 0.130 | 0.663 | 0.199 |
| **Women (n=437)** |  |  |  |  |
| Waist circumference | 0.683 (0.607-0.758) | - |  |  |
| Waist-to-hip ratio | 0.652 (0.570-0.735) | 0.429 | - |  |
| Waist-to-height ratio | 0.693 (0.619-0.767) | 0.351 | 0.289 | - |
| Body mass index | 0.691 (0.620-0.762) | 0.721 | 0.473 | 0.944 |

Values are area under the Receiver Operating Curve (AUC) and 95% confidence interval (95%CI) and P-values for the difference in the AUC between adiposity markers. WC, waist circumference; WHR, waist-to-hip ratio; WHtR, waist-to-height ratio; BMI, body mass index.

## References

1. Saeedi P, Petersohn I, Salpea P, Malanda B, Karuranga S, Unwin N, Colagiuri S, Guariguata L, Motala AA, Ogurtsova K, Shaw JE, Bright D, Williams R, Committee IDFDA. Global and regional diabetes prevalence estimates for 2019 and projections for 2030 and 2045: Results from the International Diabetes Federation Diabetes Atlas, 9(th) edition. Diabetes Res Clin Pract 2019;157:107843

2. STATS-SA. Mortality and causes of death in South Africa, 2016: Findings from death notification. Statistics South Africa, 2018, p. 1–142

3. Okosun IS, Cooper RS, Rotimi CN, Osotimehin B, Forrester T. Association of waist circumference with risk of hypertension and type 2 diabetes in Nigerians, Jamaicans, and African-Americans. Diabetes Care 1998;21:1836–1842

4. Ross R, Neeland IJ, Yamashita S, Shai I, Seidell J, Magni P, Santos RD, Arsenault B, Cuevas A, Hu FB, Griffin BA, Zambon A, Barter P, Fruchart JC, Eckel RH, Matsuzawa Y, Despres JP. Waist circumference as a vital sign in clinical practice: a Consensus Statement from the IAS and ICCR Working Group on Visceral Obesity. Nat Rev Endocrinol 2020;16:177–189

5. Pi-Sunyer FX. Obesity: criteria and classification. Proc Nutr Soc 2000;59:505–509

6. Alberti KG, Eckel RH, Grundy SM, Zimmet PZ, Cleeman JI, Donato KA, Fruchart JC, James WP, Loria CM, Smith SC, Jr., International Diabetes Federation Task Force on E, Prevention, Hational Heart L, Blood I, American Heart A, World Heart F, International Atherosclerosis S, International Association for the Study of O. Harmonizing the metabolic syndrome: a joint interim statement of the International Diabetes Federation Task Force on Epidemiology and Prevention; National Heart, Lung, and Blood Institute; American Heart Association; World Heart Federation; International Atherosclerosis Society; and International Association for the Study of Obesity. Circulation 2009;120:1640–1645

7. Mtintsilana A, Micklesfield LK, Chorell E, Olsson T, Goedecke JH. Fat redistribution and accumulation of visceral adipose tissue predicts type 2 diabetes risk in middle-aged black South African women: a 13-year longitudinal study. Nutr Diabetes 2019;9:12

8. Chantler S, Dickie K, Micklesfield LK, Goedecke JH. Longitudinal Changes in Body Fat and Its Distribution in Relation to Cardiometabolic Risk in Black South African Women. Metab Syndr Relat Disord 2015;13:381–388

9. Goedecke JH, Levitt NS, Lambert EV, Utzschneider KM, Faulenbach MV, Dave JA, West S, Victor H, Evans J, Olsson T, Walker BR, Seckl JR, Kahn SE. Differential effects of abdominal adipose tissue distribution on insulin sensitivity in black and white South African women. Obesity (Silver Spring) 2009;17:1506–1512

10. Keswell D, Tootla M, Goedecke JH. Associations between body fat distribution, insulin resistance and dyslipidaemia in black and white South African women. Cardiovasc J Afr 2016;27:177–183

11. Sumner AE, Micklesfield LK, Ricks M, Tambay AV, Avila NA, Thomas F, Lambert EV, Levitt NS, Evans J, Rotimi CN, Tulloch-Reid MK, Goedecke JH. Waist circumference, BMI, and visceral adipose tissue in white women and women of African descent. Obesity (Silver Spring) 2011;19:671–674

12. World Health Organisation. Waist circumference and waist-hip ratio: report of a WHO expert Consulatation. Geneva, 2011

13. Matsha TE, Hassan MS, Hon GM, Soita DJ, Kengne AP, Erasmus RT. Derivation and validation of a waist circumference optimal cutoff for diagnosing metabolic syndrome in a South African mixed ancestry population. Int J Cardiol 2013;168:2954–2955

14. Crowther NJ, Norris SA. The current waist circumference cut point used for the diagnosis of metabolic syndrome in sub-Saharan African women is not appropriate. PLoS One 2012;7:e48883

15. Ekoru K, Murphy GAV, Young EH, Delisle H, Jerome CS, Assah F, Longo-Mbenza B, Nzambi JPD, On’Kin JBK, Buntix F, Muyer MC, Christensen DL, Wesseh CS, Sabir A, Okafor C, Gezawa ID, Puepet F, Enang O, Raimi T, Ohwovoriole E, Oladapo OO, Bovet P, Mollentze W, Unwin N, Gray WK, Walker R, Agoudavi K, Siziya S, Chifamba J, Njelekela M, Fourie CM, Kruger S, Schutte AE, Walsh C, Gareta D, Kamali A, Seeley J, Norris SA, Crowther NJ, Pillay D, Kaleebu P, Motala AA, Sandhu MS. Deriving an optimal threshold of waist circumference for detecting cardiometabolic risk in sub-Saharan Africa. Int J Obes (Lond) 2017;

16. Motala AA, Esterhuizen T, Pirie FJ, Omar MA. The prevalence of metabolic syndrome and determination of the optimal waist circumference cutoff points in a rural South african community. Diabetes Care 2011;34:1032–1037

17. Ali SA, Soo C, Agongo G, Alberts M, Amenga-Etego L, Boua RP, Choudhury A, Crowther NJ, Depuur C, Gomez-Olive FX, Guiraud I, Haregu TN, Hazelhurst S, Kahn K, Khayeka-Wandabwa C, Kyobutungi C, Lombard Z, Mashinya F, Micklesfield L, Mohamed SF, Mukomana F, Nakanabo-Diallo S, Natama HM, Ngomi N, Nonterah EA, Norris SA, Oduro AR, Some AM, Sorgho H, Tindana P, Tinto H, Tollman S, Twine R, Wade A, Sankoh O, Ramsay M. Genomic and environmental risk factors for cardiometabolic diseases in Africa: methods used for Phase 1 of the AWI-Gen population cross-sectional study. Glob Health Action 2018;11:1507133

18. Ramsay M, Crowther N, Tambo E, Agongo G, Baloyi V, Dikotope S, Gomez-Olive X, Jaff N, Sorgho H, Wagner R, Khayeka-Wandabwa C, Choudhury A, Hazelhurst S, Kahn K, Lombard Z, Mukomana F, Soo C, Soodyall H, Wade A, Afolabi S, Agorinya I, Amenga-Etego L, Ali SA, Bognini JD, Boua RP, Debpuur C, Diallo S, Fato E, Kazienga A, Konkobo SZ, Kouraogo PM, Mashinya F, Micklesfield L, Nakanabo-Diallo S, Njamwea B, Nonterah E, Ouedraogo S, Pillay V, Somande AM, Tindana P, Twine R, Alberts M, Kyobutungi C, Norris SA, Oduro AR, Tinto H, Tollman S, Sankoh O. H3Africa AWI-Gen Collaborative Centre: a resource to study the interplay between genomic and environmental risk factors for cardiometabolic diseases in four sub-Saharan African countries. Glob Health Epidemiol Genom 2016;1:e20

19. Friedewald WT, Levy RI, Fredrickson DS. Estimation of the Concentration of Low-Density Lipoprotein Cholesterol in Plasma, Without Use of the Preparative Ultracentrifuge. Clinical Chemistry 1972;18:499–502

20. World Health Organization,International Diabetes F. Definition and diagnosis of diabetes mellitus and intermediate hyperglycaemia : report of a WHO/IDF consultation. Geneva, World Health Organization, 2006.

21. Youden WJ. Index for rating diagnostic tests. Cancer 3:4

22. Bewick V, Cheek L, Ball J. Statistics review 13: receiver operating characteristic curves. Crit Care 2004;8:508–512

23. Peer N, Steyn K, Levitt N. Differential obesity indices identify the metabolic syndrome in Black men and women in Cape Town: the CRIBSA study. J Public Health (Oxf) 2016;38:175–182

24. National Department of Health, ICF. South Africa Demographic and Health Survey 2016. Pretoria, National Department of Health -NDoH - ICF, 2019

25. Kabakambira JD, Baker RL, Jr., Briker SM, Courville AB, Mabundo LS, DuBose CW, Chung ST, Eckel RH, Sumner AE. Do current guidelines for waist circumference apply to black Africans? Prediction of insulin resistance by waist circumference among Africans living in America. BMJ Glob Health 2018;3:e001057

26. Woudberg NJ, Goedecke JH, Blackhurst D, Frias M, James R, Opie LH, Lecour S. Association between ethnicity and obesity with high-density lipoprotein (HDL) function and subclass distribution. Lipids Health Dis 2016;15:92

27. Woudberg NJ, Lecour S, Goedecke JH. HDL Subclass Distribution Shifts with Increasing Central Adiposity. Journal of Obesity 2019;2019:1–6

28. Greiner R, Nyrienda M, Rodgers L, Asiki G, Banda L, Shields B, Hattersley A, Crampin A, Newton R, Jones A. Associations between low HDL, sex and cardiovascular risk markers are substantially different in sub-Saharan Africa and the UK: analysis of four population studies. BMJ Glob Health 2021;V6

29. Knight MG, Goedecke JH, Ricks M, Evans J, Levitt NS, Tulloch-Reid MK, Sumner AE. The TG/HDL-C ratio does not predict insulin resistance in overweight women of African descent: a study of South African, African American and West African women. Ethn Dis 2011;21:490–494

30. Sumner AE, Cowie CC. Ethnic differences in the ability of triglyceride levels to identify insulin resistance. Atherosclerosis 2008;196:696–703

31. Sumner AE, Zhou J, Doumatey A, Imoisili OE, Amoah A, Acheampong J, Oli J, Johnson T, Adebamowo C, Rotimi CN. Low HDL-Cholesterol with Normal Triglyceride Levels is the Most Common Lipid Pattern in West Africans and African Americans with Metabolic Syndrome: Implications for Cardiovascular Disease Prevention. CVD Prev Control 2010;5:75–80

32. Sumner AE. Ethnic differences in triglyceride levels and high-density lipoprotein lead to underdiagnosis of the metabolic syndrome in black children and adults. J Pediatr 2009;155:S7 e7–11

33. Yu SS, Ramsey NL, Castillo DC, Ricks M, Sumner AE. Triglyceride-based screening tests fail to recognize cardiometabolic disease in African immigrant and African-American men. Metab Syndr Relat Disord 2013;11:15–20

34. Evans J, Micklesfield L, Jennings C, Levitt NS, Lambert EV, Olsson T, Goedecke JH. Diagnostic ability of obesity measures to identify metabolic risk factors in South African women. Metab Syndr Relat Disord 2011;9:353–360

35. Kengne AP, Erasmus RT, Levitt NS, Matsha TE. Alternative indices of glucose homeostasis as biochemical diagnostic tests for abnormal glucose tolerance in an African setting. Prim Care Diabetes 2017;11:119–131

36. Wade AN, Crowther NJ, Abrahams-Gessel S, Berkman L, George JA, Gomez-Olive FX, Manne-Goehler J, Salomon JA, Wagner RG, Gaziano TA, Tollman SM, Cappola AR. Concordance between fasting plasma glucose and HbA1c in the diagnosis of diabetes in black South African adults: a cross-sectional study. BMJ Open 2021;11:e046060

37. Peer N, Steyn K, Lombard C, Lambert EV, Vythilingum B, Levitt NS. Rising diabetes prevalence among urban-dwelling black South Africans. PLoS One 2012;7:e43336

38. Motala AA, Esterhuizen T, Gouws E, Pirie FJ, Omar MA. Diabetes and other disorders of glycemia in a rural South African community: prevalence and associated risk factors. Diabetes Care 2008;31:1783–1788

